# Data driven inference of the reproduction number (*R*_0_) for COVID-19 before and after interventions for 51 European countries

**DOI:** 10.1101/2020.05.21.20109314

**Authors:** Petr Karnakov, George Arampatzis, Ivica Kičić, Fabian Wermelinger, Daniel Wälchli, Costas Papadimitriou, Petros Koumoutsakos

**Affiliations:** Computational Science and Engineering Laboratory, ETH Zurich, Switzerland; Department of Mechanical Engineering, University of Thessaly, Greece

**Keywords:** COVID-19, Bayesian inference, SIR model, interventions

## Abstract

The reproduction number (*R*_0_) is broadly considered as a key indicator for the spreading of the COVID-19 pandemic. The estimation of its value with respect to the key threshold of 1.0 is a measure of the need, and eventually effectiveness, of interventions imposed in various countries. Here we present an online tool for the data driven inference and quantification of uncertainties for *R*_0_ as well as the time points of interventions for 51 European countries. The study relies on the Bayesian calibration of the simple and well established SIR model with data from reported daily infections. The model is able to fit the data for most countries without individual tuning of parameters. We deploy an open source Bayesian inference framework and efficient sampling algorithms to present a publicly available GUI cse-lab.ethz.ch/coronavirus that allows the user to assess custom data and compare predictions for pairs of European countries. The results provide a ranking based on the rate of the disease’s spread suggesting a metric for the effectiveness of social distancing measures. They also serve to demonstrate how geographic proximity and related times of interventions can lead to similarities in the progression of the epidemic.

## 1 Introduction

The forecasting of the evolution of the COVID-19 pandemic and the effects of lock-downs of any kind as well as social distancing measures are critical components for decision makers across the world. There is a broad range of data analysis tools and forecasting models that have been deployed since the beginning of 2020 to assess the spread of the disease as well as the expected number of infections and numbers of deaths [1, 2, 3]. A metric that is often deployed to quantify the progress of the disease is the reproduction number (*R*_0_). While *R*_0_ exhibits significant complexity [4], it is broadly considered that values of *R*_0_ above 1.0 indicate a rapid expansion of the infections. Governments have resorted to impose social distancing measures to reduce this number well below 1.0. The estimation of *R*_0_ hinges on the forecasting model and the data used to infer their parameters. Moreover, an often overlooked fact is that, the method by which inference is performed also plays a role on these estimates. A well established technique for such inference is Bayesian inference [5]. However, even though Bayesian inference is a very potent method to estimate uncertainties in model structure and parameters, model parameters may be unidentifiable for the chosen type and amount of data as well as the choice of priors [6, 7].

In this work we deploy Bayesian inference to quantify the evolution of *R*_0_ as well as the time points of interventions for 51 European countries. The study relies on the Bayesian calibration of the simple, and well established SIR model [8], extended to account for interventions, with data from reported daily infections. We present an online interface that allows for entry of customized data and comparisons between countries. The parameters of the model include the reproduction number, the day of the first intervention and a reduction factor for the reproduction number. By inferring these parameters from data, we identify when interventions become effective and determine the reproduction number before and after the measures have become effective. This indicates how well the imposed restrictions are able to slow down the spread of the disease.

We remark that very recently a related work [9] inferred the intervention points and the evolution of *R*_0_ in Germany. We note that in the present work we apply our model to 51 European countries. Our model is able to fit the data for the majority of the countries without individual tuning of the parameters or the inference method for each country. We note that our results are in excellent agreement with the work previously mentioned [9] for Germany. Moreover our results indicate that capturing accurately the results for one country does not generalize well with the same accuracy to all countries. We visualize the inferred quantities on a map and rank the countries by the disease’s spread rate and the effectiveness of the interventions. We find that the results of our model are consistent with those from related studies in Switzerland [10]. The results suggest that at the current stage of the pandemic, countries can be categorized according to the inferred *R*_0_. Most countries have imposed their restrictions within the first weeks of the epidemics starting March 2020, and since then about 80% of them have managed to bring their country specific *R*_0_ below 1 which indicates a decaying epidemic. Other countries, such as Poland, Sweden and Moldova, remain at a stage with larger *R*_0_.

## 2 Methods

### 2.1 Data

We calibrate the well established SIR model using data from daily confirmed cases reported in the open source repository Humanistic GIS Lab, University of Washington [11], whose main data source for European countries is the World Health Organization [12]. To avoid the stochastic regime of the system, for each country we consider data only after the number of confirmed cases exceeds a total of five and a total of two per million of population. Population figures for countries are retrieved from the open repository [13]. The analysis in this paper considers data up to May 18, 2020. Official times of interventions that we use for comparison in Section 3.2 are taken from [14] which aggregates the official announcements of social distancing measures by various governments.

### 2.2 Epidemic modeling

The SIR model describes the evolution of susceptible (S), infected (I), and removed (R) population,

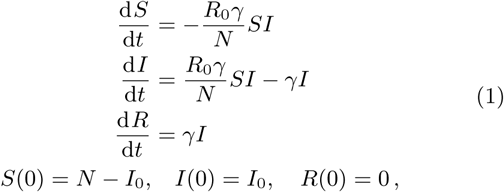

where *R*_0_ is the reproduction number, *γ* is the removal rate (including recovery and mortality), *I*_0_ is the initial number of infected individuals and *N* is the size of the population. To model an intervention or a series of measures taken by the government, we consider *R*_0_ to be a piece-wise linear function of time split into three phases,

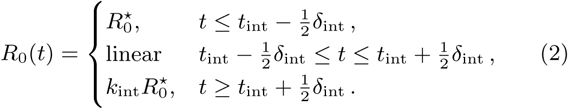

The three phases are: (i) uncontrolled disease outbreak before the first intervention takes place (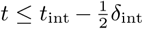), (ii) an adoption phase during which one or multiple measures take place (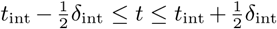), and (iii) the period after all interventions have become effective (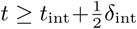). In the first and last regime we assume that the reproduction number *R*_0_(*t*) remains constant, and during the adoption phase we assume a linear transition from 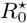 to 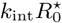. The intervention time *t*_int_ and the reduction factor *k*_int_ ∊ (0, 1) are inferred from the data, while the duration of transition remains fixed at *δ*_int_ = 10 days.

### 2.3 Bayesian inference

We quantify the uncertainty in the extended SIR model as described above using data of daily reported infected people of each country. We denote this data with 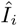 corresponding to day *t_i_*. From the SIR model, the daily incidence is computed

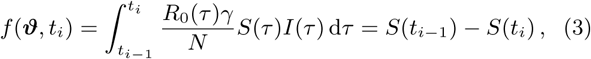

where 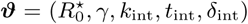 are the model parameters of the SIR model including an intervention as described above. The initial number of infected individuals is set from the available data so that 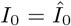. We consider the following generative model for the daily incidence data

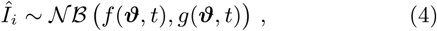

where 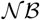is the negative Binomial parametrized by the mean *f*(*ϑ*, *t*) and the dispersion *g*(*ϑ*, *t*). Notice that using this parametrization, the variance of an observation 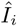 is given by *f*(*ϑ*, *t_i_*) + *f(ϑ, t_i_*)^2^/*g(ϑ, t_i_)*. Here, the dispersion function is constant, i.e. *g*(*ϑ*, *t*) = *r*, where the parameter vector has been extended to include the dispersion parameter 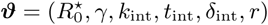.

We denote the collection of observations by the vector 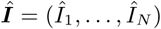 and the corresponding observation times by 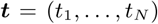. For the estimation of the probability of *ϑ* conditioned on the data, we apply Bayes’ theorem,

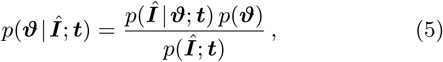

where *p*(***Î*** | *ϑ*; *t*) is the likelihood function, *p*(*ϑ*) is the prior probability distribution and *p*(***Î***; *t*) is the model evidence.

Assuming the observations are independent, the likelihood function is given by

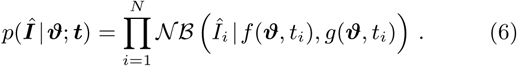

We assume uninformative priors for the model parameter 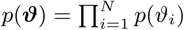. The lower and upper bounds of the priors are summarized in Table 1. We assume that the removal rate *γ* is the same in all countries and set it to *γ* = 1/5.2*l*days following [15].

**Table 1:** Parameters and the prior distributions for the model discussed in Section 2.2.

| Parameter | Type | Value | Unit |
| --- | --- | --- | --- |
| $R_0^*$ | Inferred | Uniform(1, 4) | - |
| $\gamma$ | Fixed | 1/5.2 [15] | 1/days |
| $k_{\text{int}}$ | Inferred | Uniform(0.1, 10) | - |
| $t_{\text{int}}$ | Inferred | Uniform(0, 80) | days |
| $\delta_{\text{int}}$ | Fixed | 10 | days |
| $r$ | Inferred | Uniform(0, 50) | - |

The inference of *ϑ* is performed using Korali [16, 17], a high-performance framework for uncertainty quantification and optimization of computational models. The posterior distribution was estimated with Bayesian Annealed Sequential Importance Sampling (BASIS), a reduced variant of the Transitional Markov Chain Monte Carlo (TMCMC) algorithm [18], ran with 5000 samples and default parameters taken from [19].

### 2.4 Graphical User Interface

We provide an online interface for real time evaluation of our predictive model^1^. The interface allows for side-by-side comparison of two countries with default input data obtained from [11]. Modification of individual data points or substitution of different data sets can be achieved by changing the data in the input form. The accuracy of the Bayesian inference can be improved by increasing the number of samples used by the solver. User requests are forwarded from the web server to a remote workstation where the runs are executed. The back-end code of our model evaluation is implemented in the Python, while performance critical parts are implemented in C++. The mean and confidence interval of the computed prediction for total and new daily infected cases are displayed together with input data. The inferred intervention date, *t*_int_, is highlighted by the vertical line with corresponding reproduction number before and after the intervention. Figure 1 shows a screenshot of our online interface comparing the predictions for Switzerland and Sweden. Social distancing measures introduced by the Swiss government starting on March 17, 2020 have been more successful in stopping the epidemic, compared to the less strict approach of Sweden. The samples drawn from the posterior distribution are further shown in the columns of the selected countries.

**Figure 1:**
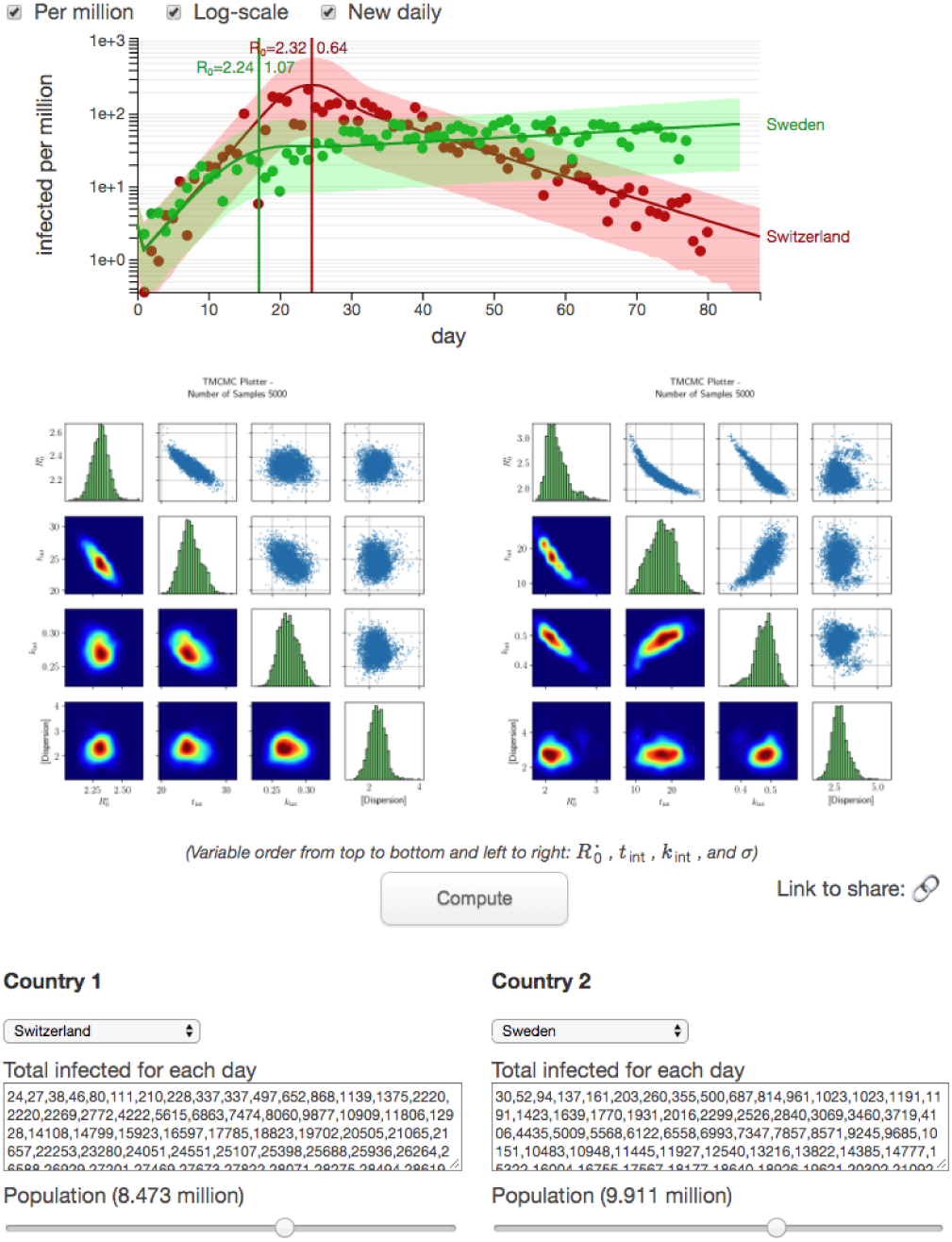
Screenshot of the online interface comparing predictions for Switzerland and Sweden on May 21 with 5000 samples https://www.cse-lab.ethz.ch/coronavirus/.

## 3 Results and Discussion

### 3.1 Verification

We verify the presented intervention model using the results of a recent related study [9] that reported the results of a similar SIR model with interventions. They also performed Bayesian inference of the model parameters applied to Germany. That model has more parameters to describe multiple interventions and considers a periodic modulation of the daily reported cases to account for trends such as under-reporting on the weekends. We compare our results to this study in Fig. 4 where the reported quantities are the effective growth rate *λ*\*(*t*) = *γ(R*_0_(*t*) − 1), the number of new daily cases and the total number of cases. The effective growth rate relates to one limiting case of the SIR model where the total number of infected individuals is small compared to the population such that *S/N* → 1 and the model reduces to an exponential growth *I(t*) ∝ exp(*λ*\**t*), which is valid at early stages of the epidemic. Fig. 5 shows the corresponding parameters drawn from the posterior distribution.

We want to highlight that our model is simpler and has fewer parameters than the one suggested in [9]. Also we observe that the fitted model agrees well with the reported data and provides an estimate for the intervention time for Germany.

### 3.2 Application to 51 European countries

We apply our framework to 51 countries of the European continent and infer the model parameters for each country separately. The inferred reproduction number before and after intervention is shown in Fig. 2. The map and the scatter plot reveal the differences in the values of *R*_0_ and effects of interventions among various countries. Certain countries show strong similarity in the inferred parameters. For example, the neighboring Switzerland and Austria both have *R*_0_ dropping from 2.3 to 0.7 after interventions, even though the latter imposed more constraining measures. The same is evident for other pairs such as Italy and France and also Poland and Ukraine. However, no single explanation for these similarities and differences can be suggested. Physical proximity of the countries does not always lead to similar progression of the epidemic. One example is Poland where the epidemic has started slower but the imposed restrictions have proved less efficient than in adjacent Lithuania. The disease’s development depends on the types of measures taken by the governments as well as the demographics and the particular features of the social interactions in the population. For instance, the UK and Sweden initially decided against imposing strict measures and therefore undergo a longer epidemic with more infections than their neighbors, Ireland and Norway, which corresponds to larger inferred values of *R*_0_.

**Figure 2:**
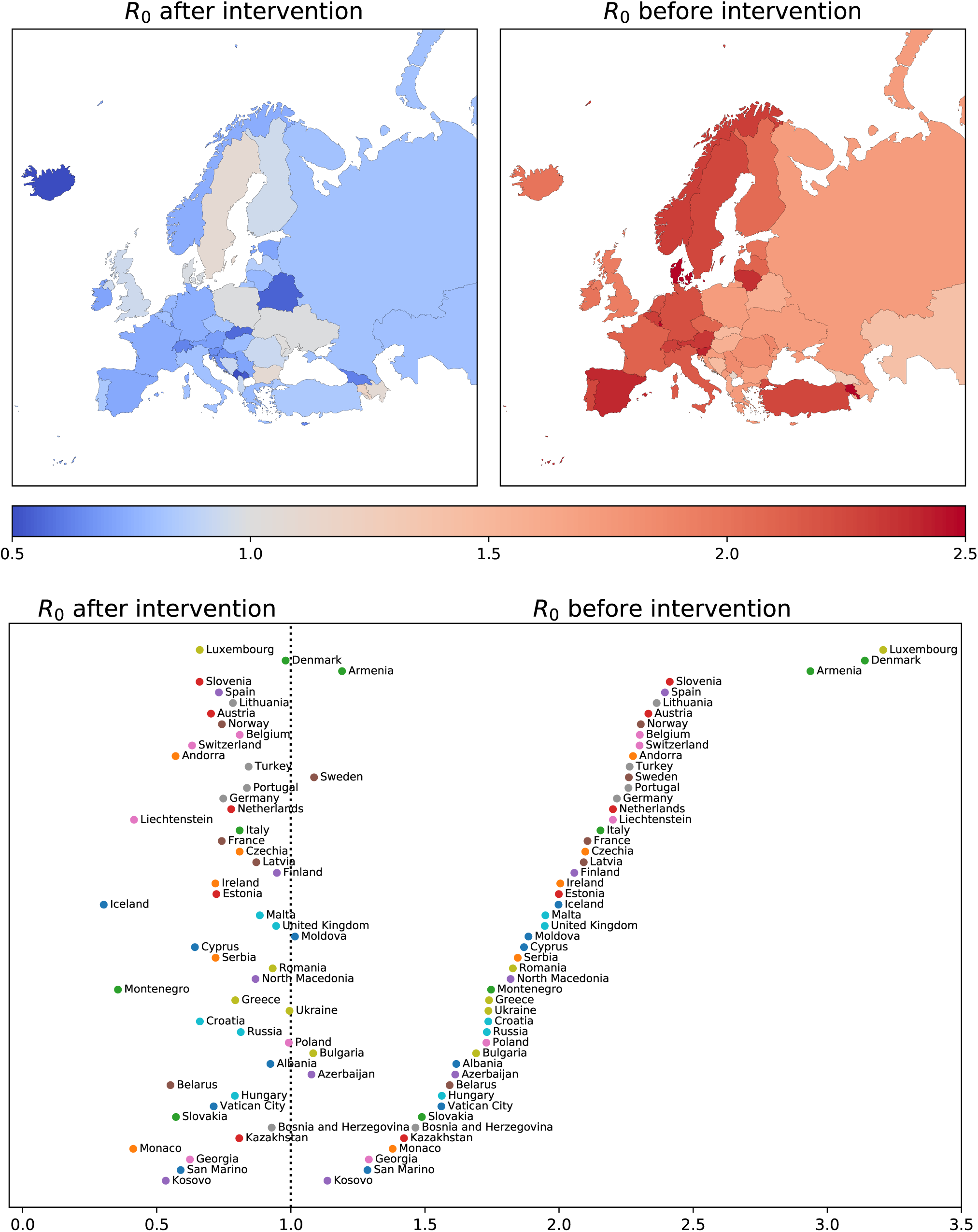
Inferred mean of the reproduction number *R*_0_ before (right) and after intervention (left) indicated in color on a map of Europe [20] (top) and in a scatter plot (bottom), where the countries are ordered by the value of *R*_0_ before interventions.

Another inferred parameter is the intervention time. Figure 3 shows the relation between the reproduction number and the intervention time measured from the beginning of the epidemic in each country. One trend is apparent from the values of *R*_0_ before intervention: countries with larger initial *R*_0_ tend to introduce the restrictions earlier.

**Figure 3:**
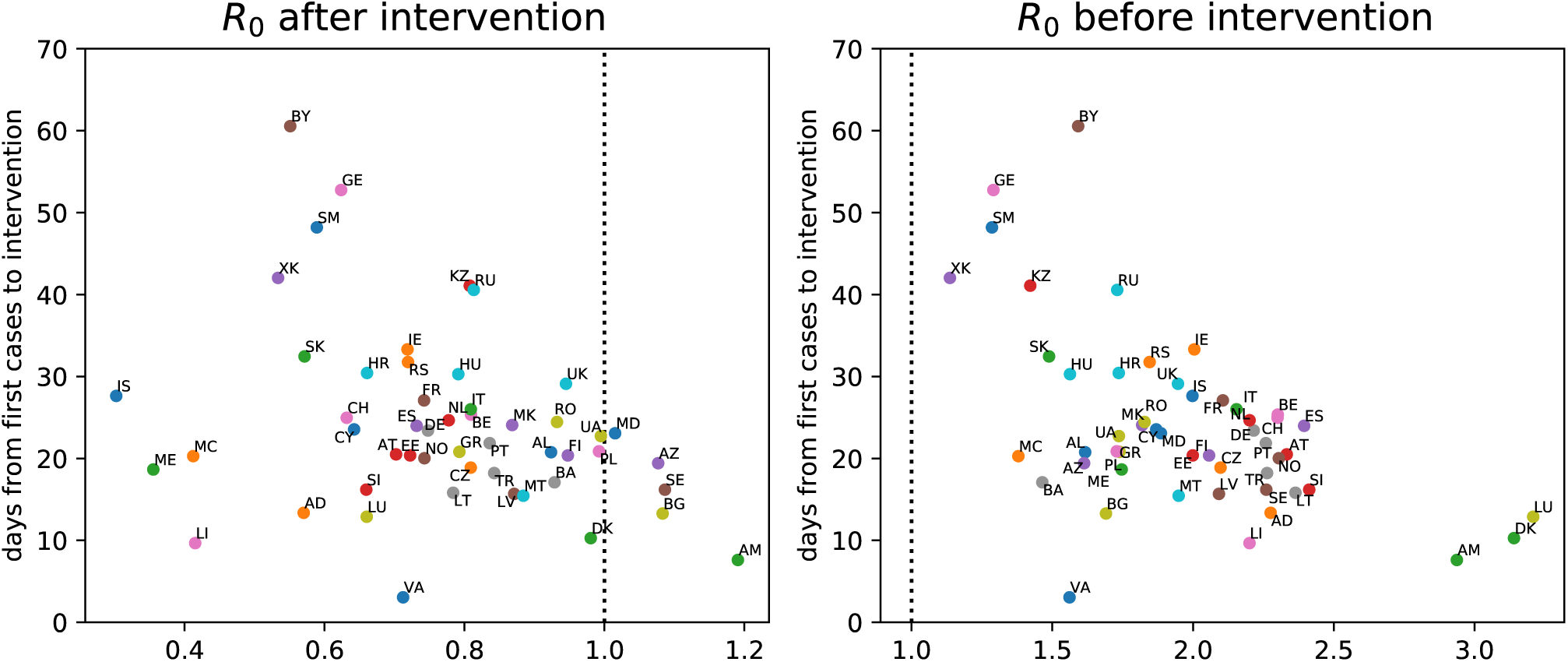
Inferred mean of the reproduction number *R*_0_ before (right) and after intervention (left) against the intervention time measured from the beginning of the epidemic in each country. The colors of dots correspond to Fig. 2.

**Figure 4:**
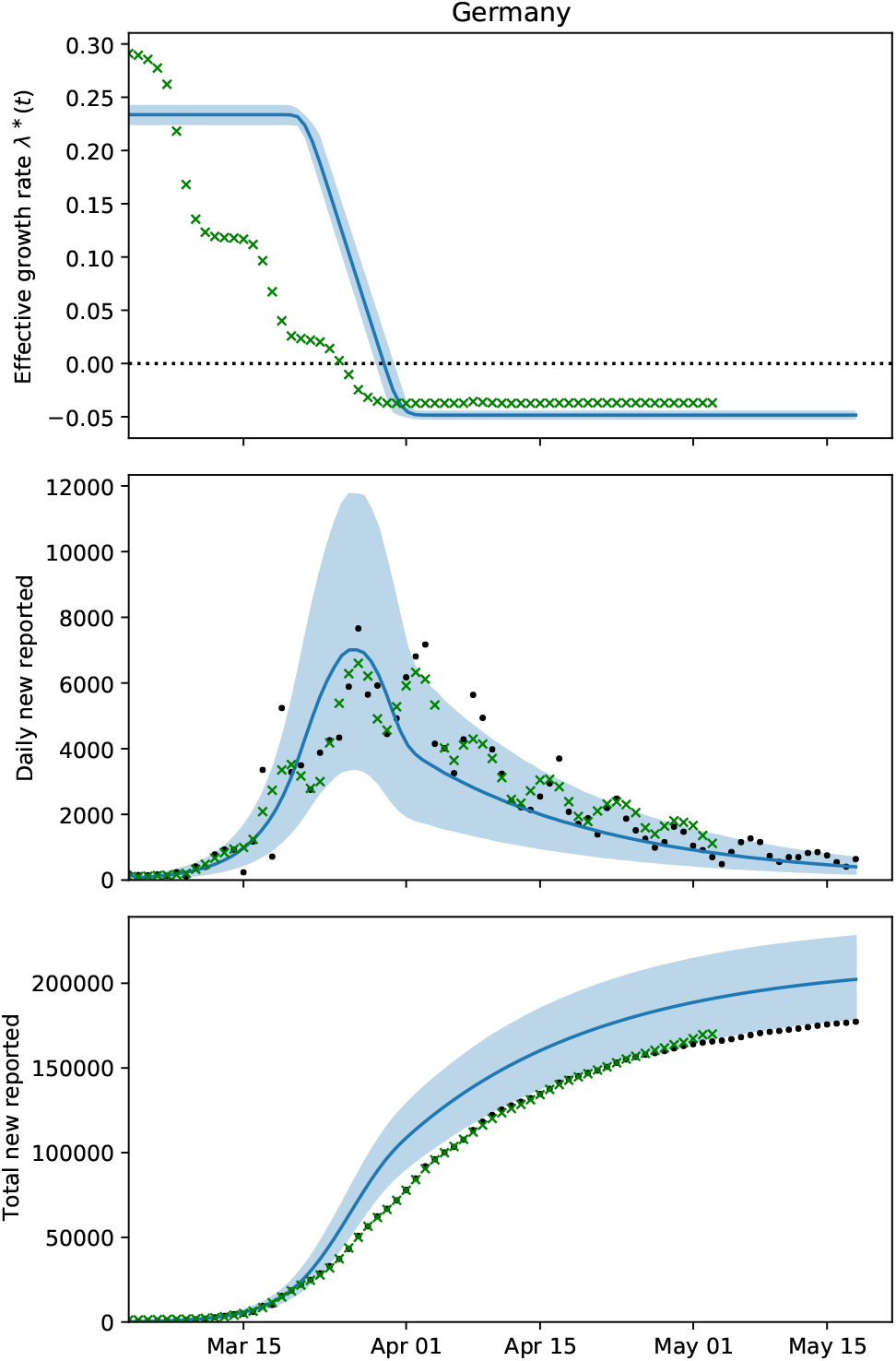
The effective growth rate (top), the number of new infections per day (middle) and the total number of infections in Germany (bottom) fitted by present model (blue lines and shades) compared to median predictions from [9] (green crosses). The solid lines show the mean prediction and the shades are the 90% confidence intervals.

**Figure 5:**
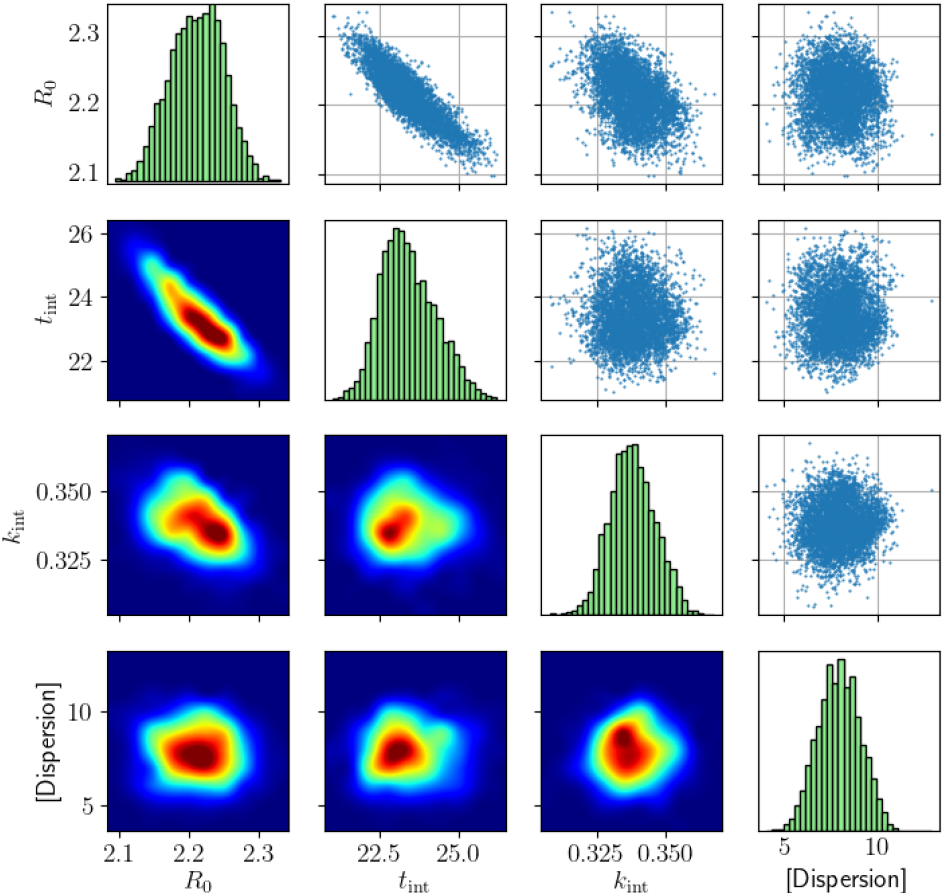
Inferred parameters (the initial reproduction number 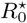, intervention time *t*_int_, reduction factor after intervention *k*_int_, and the dispersion parameter *r* of the negative binomial distribution (Eq. (4))) for Germany: samples drawn from the posterior distribution (upper triangle), marginal distributions of the individual parameter (diagonal) and likelihood heat map (lower triangle).

The inferred intervention times are shown in Fig. 6 and listed in Table 2 together with the times when the restrictions have been officially announced. The countries are ordered by the inferred intervention time which helps to identify countries with similar inferred parameters. For certain countries, such as Switzerland, Germany and Greece, the inferred intervention time matches the official announcements with a delay of at most one week. In other cases, the delays can reach up to one month. This may indicate that the measures are imposed gradually and do not show an immediate effect.

**Table 2:** Inferred mean intervention time compared to the official beginning of social distancing measures [14] in each country.

| Country | Inferred | Official | Inferred – Official |
| --- | --- | --- | --- |
| Albania | 2020-03-31 | 2020-03-13 | +18 |
| Andorra | 2020-03-29 |  |  |
| Armenia | 2020-03-20 | 2020-03-24 | −3 |
| Austria | 2020-03-23 | 2020-03-16 | +7 |
| Azerbaijan | 2020-04-03 | 2020-03-31 | +3 |
| Belarus | 2020-05-13 |  |  |
| Belgium | 2020-03-29 | 2020-03-18 | +11 |
| Bosnia and Herzegovina | 2020-04-06 |  |  |
| Bulgaria | 2020-03-25 |  |  |
| Croatia | 2020-04-02 | 2020-03-18 | +15 |
| Cyprus | 2020-04-04 |  |  |
| Czechia | 2020-03-27 |  |  |
| Denmark | 2020-03-14 | 2020-03-11 | +3 |
| Estonia | 2020-03-26 |  |  |
| Finland | 2020-03-25 | 2020-03-27 | −1 |
| France | 2020-03-28 | 2020-03-17 | +11 |
| Georgia | 2020-04-28 | 2020-03-31 | +28 |
| Germany | 2020-03-26 | 2020-03-23 | +3 |
| Greece | 2020-03-26 | 2020-03-23 | +3 |
| Hungary | 2020-04-13 | 2020-03-28 | +16 |
| Iceland | 2020-03-30 |  |  |
| Ireland | 2020-04-07 | 2020-03-12 | +26 |
| Italy | 2020-03-20 | 2020-03-09 | +11 |
| Kazakhstan | 2020-04-29 |  |  |
| Kosovo | 2020-05-06 | 2020-03-14 | +53 |
| Latvia | 2020-03-25 |  |  |
| Liechtenstein | 2020-03-26 |  |  |
| Lithuania | 2020-03-29 | 2020-03-16 | +13 |
| Luxembourg | 2020-03-24 | 2020-03-18 | +6 |
| Malta | 2020-03-26 |  |  |
| Moldova | 2020-04-06 |  |  |
| Monaco | 2020-04-04 |  |  |
| Montenegro | 2020-04-07 | 2020-03-24 | +14 |
| Netherlands | 2020-03-29 | 2020-03-16 | +13 |
| North Macedonia | 2020-04-03 |  |  |
| Norway | 2020-03-20 | 2020-03-12 | +8 |
| Poland | 2020-04-04 | 2020-03-13 | +22 |
| Portugal | 2020-03-30 | 2020-03-19 | +11 |
| Romania | 2020-04-04 | 2020-03-25 | +10 |
| Russia | 2020-05-01 | 2020-03-30 | +32 |
| San Marino | 2020-04-21 | 2020-03-14 | +38 |
| Serbia | 2020-04-12 | 2020-03-15 | +28 |
| Slovakia | 2020-04-13 | 2020-03-16 | +28 |
| Slovenia | 2020-03-22 |  |  |
| Spain | 2020-03-26 | 2020-03-14 | +12 |
| Sweden | 2020-03-19 |  |  |
| Switzerland | 2020-03-25 | 2020-03-17 | +8 |
| Turkey | 2020-04-06 | 2020-04-23 | −16 |
| Ukraine | 2020-04-16 | 2020-03-17 | +30 |
| United Kingdom | 2020-04-04 | 2020-03-23 | +12 |
| Vatican City | 2020-04-01 |  |  |

The present code and GUI are open source and can be adapted to various countries and data. We believe that the present study indicates the power and need for data driven Bayesian inference in the forecasting of the evolution of COVID-19. We note that the described methodology is readily extensible to other epidemic models and types of data that are the subject of our ongoing studies.

**Figure 6:**
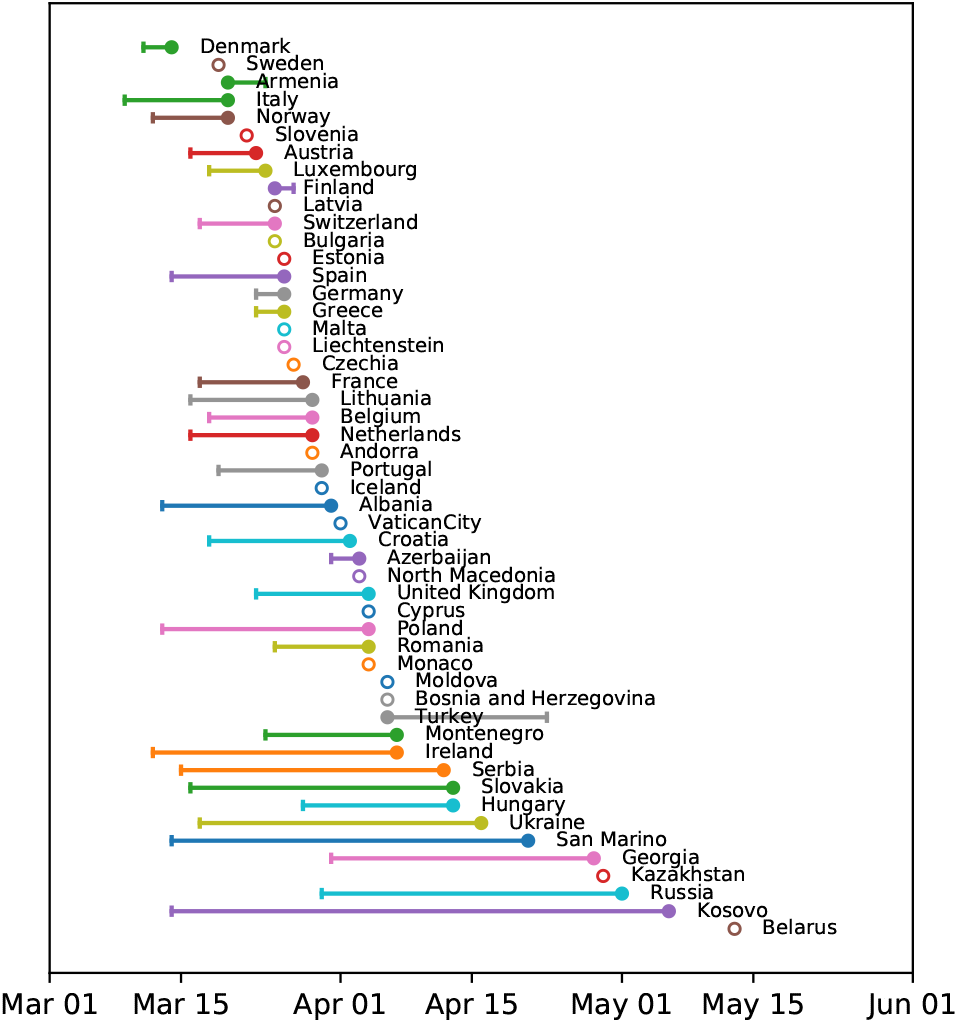
Inferred mean intervention time (circles) compared to the official beginning (vertical bars) of social distancing measures [14] in each country. Empty circles indicate missing data for the official time. The countries are ordered by the inferred intervention time such that countries with similar parameters are located closer.

### 3.3 Disclosure statement

No financial support and no other potential conflict of interest relevant to this article were reported.

## Data Availability

All data used in the manuscript is publicly available.

https://github.com/samayo/country-json

https://github.com/jakobzhao/virus

http://naciscdn.org/naturalearth/packages/natural_earth_vector.zip

## 3.4 Acknowledgments

We acknowledge valuable discussions with Lucas Amoudruz, Martin Boden, Michalis Chatzimanolakis and Pascal Weber (ETHZ). Sergio Martin (ETHZ) provided valuable technical assistance with the Korali framework. We acknowledge funding by ETH Zürich and computing resources by the Swiss Supercomputing center (CSCS).

## Footnotes

1 https://www.cse-lab.ethz.ch/coronavirus/

